# Differences of clinical and imaging findings in multiple generations of secondary COVID-19 infection in Xi’an, China

**DOI:** 10.1101/2020.05.23.20109496

**Authors:** Runqing Li, Zhijie Jian, Chao Jin, Yan Wang, Ting Liang, Zhe Liu, Huifang Zhao, Zekun Wang, Jie Zhou, Lingxia Zeng, Jian Yang

## Abstract

**PURPOSE:** In the global presence of secondary infections with the coronavirus disease 2019 (COVID-19), little is known about the transmission characteristics of COVID-19 outside Wuhan, China. We evaluated differences in clinic and radiologic findings of multiple generations of COVID-19 infection in Xi’an (Shaanxi, China) to provide more clues for the correct estimate of the disease.

**METHODS:** All COVID-19 infected patients reported in Xi’an up to 10 February 2020 were included for this analysis. Among these cases, clinical and chest CT data of 62 cases were obtained from three hospital in Xi’an. With this information, patients were grouped on basis of exposure history and transmission chains as first-generation, second-generation and third-generation patients. We described clinical characteristics and evaluated CT score/patterns in these COVID-19 cases.

**RESULTS:** There was a clear age differences in multiple generations with COVID-19 infection. Above two thirds of the second-generation (75.0%) and third-generation patients (77.8%) were aged ≥45 years while 40.0% of first-generation cases at this age (*p*=0.001). More than half of second-generation patients (52.8%) and third-generation patients (55.6%) have comorbidities and is predominantly hypertensive (22.8% of second-generation vs. 27.8% of third-generation infections). The main exposure of second- and third-generation patients in Xi’an is family exposure (35.2%). For evaluation of CT findings of pulmonary involvement, the total CT score were 4.22±3.00 in first-generation group, 4.35±3.03 in second-generation group and 7.62±3.56 in third-generation group (*p*<0.001). In all of three generations, the predominant pattern of abnormality observed was organizing pneumonia (65.5% in first-generation group, 61.5% in second-generation group and 71.4% in third-generation group). The average courses of the disease in third-generation infections has obviously extension (22.93±7.22 days of first-generation, 21.53±8.31 days of second-generation vs. 31.00±8.12 days of third-generation group, *p*=0.004). There were no significant differences of the pulmonary sequelae among three generation patients.

**CONCLUSION:** There is more serious pulmonary infection of COVID-19 pneumonia in second- and third-generation patients, which might be attribute to the elder age and comorbidity of these patients.

## Introduction

Since its discovery in Wuhan (Hubei, China) in Nov. 2019, the coronavirus disease 2019 (COVID-19) has continued to spread rapidly to China and worldwide (1). As of April 19, the World Health Organization (WHO) has been notified of 2,241,359 laboratory-confirmed cases of COVID-19 pneumonia infection, including at least 152,551 related deaths (2). WHO therefore have made the assessment that COVID-19 can be characterized as a pandemic and seems to be more transmissible than MERS and SARS (3).

On Jan 23, 2020, Xi’an, China reported its first confirmed case of COVID-19 infection in a 33-year-old woman who has contact with people from Hubei, which was then followed by chains of secondary infections (4). As of February 10, 2020, a total of 104 COVID-19 cases have been reported in Xi’an (5). In the presence of secondary generation infections, an understanding of the dynamic transmission in COVID-19 pneumonia would contribute to effective infection prevention (6-12). Meanwhile, understanding of the main differences in clinical and radiologic characteristics of multiple generations can improve the surveillance and diagnosis of secondary infections (1).

Here, we provide an analysis of data on the multiple generations of secondary cases with COVID-19 infection in Xi’an (Shaanxi, China) to describe the epidemiologic characteristics of multiple generation infections. Additional, we explore the differences in CT findings and laboratory-testing in multiple-generation patients to improve understanding of potential significance of these data in multiple generation infections.

### Materials and Methods

This study was approved by the institutional review board (Number: XJTU1AF2020LSK-011). Informed patients’ consent was waived.

### Data Collection

In Xi’an, the first imported infection was confirmed on 23 January, 2020 (4). Then, Shaanxi province has activated first-level public health emergency response since 25 January, 2020, with calling on home quarantine, traffic control and suspension of public places. In this study, we collected information of 99 cases who have basic demographic information, date of symptom onset, medical history and epidemiological information, and 62 among them with additional information including symptoms, chest CT scans, laboratory findings, as well as treatments and outcomes data. Two researchers independently double checked the data collected by reviewing the data collection forms.

### Case Definition

All the patients were diagnosed based on the 7^th^ edition preliminary diagnosis and treatment protocols for COVID-19 from the National Health Commission of the People’s Republic of China (13). The date of symptom onset was defined as the first day when the symptom was noticed.

According to the exposure history, the patients were divided into three groups: first-generation group, second-generation group and third-generation group (14). The first-generation standard was as follow: patients clearly relevant to Wuhan (exposed to Wuhan and/or contacted with people from Wuhan) during two weeks before symptom onset. Patients infected by the first-generation are defined as second generation. The third-generation patients were cases infected by second-generation cases (and cases with no clear contact history with patients of first-generation as well as with no relevant to Wuhan). Most individuals had one source of exposure. For the few patients with several possible contexts of infection or infectors, we defined on the basis of close exposure and/or the latest exposure (10).

All 62 patients underwent initial CT after onset of symptoms. They also underwent follow-up CT scans to evaluate the progression of the disease according to the 7th edition preliminary diagnosis and treatment protocols for COVID-19 (13). The primary clinical outcome was discharge or adverse outcome (admission to ICU, use of mechanical ventilation, ECMO or death). All the following criteria should be met for hospital discharge: (1) normal temperature lasting longer than 3 days, (2) absence of respiratory symptoms (such as absence of hypoxemia, normalization of respiratory rate or normal SpO_2_ values), (3) substantially improvement of lung lesions on chest CT images, and (4) 2 consecutively negative RT-PCR test results separated by at least 1 day. After discharge, all patients were kept under surveillance and quarantined at home for at least 14 days. 2-3 weeks after discharge, 50 cases examined pulmonary sequelae by chest CT imaging.

### Image Acquisition

Chest CT scans were performed by 16- to 64-multidector CT scanners (Philips Brilliant 16, Philips Healthcare; GE VCT LightSpeed 64, GE Healthcare; Somatom Sensation 64, Siemens Healthcare). The CT parameters were as follows: 120 kVp of tube voltage, current intelligent control (auto mA) of 30-300 mA, and slice thickness/ slice interval of 0.6–3.0 mm. All CT examinations were performed without intravenous contrast material.

### Imaging Interpretation

Two experienced radiologists reviewed and described the CT demonstrations according to peer-reviewed literature of viral pneumonia (15). For each of 62 cases, the initial CT imaging was evaluated for the following characteristics: ground glass opacities (GGO) only, consolidation only, GGO and consolidation, linear opacity only, GGO and linear opacity, consolidation and linear opacity, with three signs and negative CT of lobe.

Additionally, we categorized the CT findings into four patterns by taking previous reports (16): Negative CT (with none lesion in lung), bilateral bronchopneumonia (peribronchiolar focal lesion with GGO or consolidation), organizing pneumonia (multiple focal lesions with consolidation or GGO, and/or interlobular septal thickening predominately distributed along middle-lower and subpleural lung zone or bronchovascular bundles; or if serial CTs were scanned, increasing number and/or size of lesion with disease progression within 2 weeks after onset) and diffuse alveolar damage patterns (extensive distribution of consolidation with or without GGO diffusely in the entire lungs in the first week after onset).

The pulmonary abnormalities involvement was quantitatively estimated by a semi-quantitative scoring system. Each of the 5 lung lobes was visually scored from 0 to 4 as: 0, no involvement; 1, <25% involvement; 2, 25%-49% involvement; 3, 50%-75% involvement; 4, >75% involvement. The sum of the individual lobar scores were the total CT scores, which ranged from 0 (no involvement) to 20 (maximum involvement). Finally, pulmonary sequelae of lesion resolution was examined by chest CT imaging at 2-3 weeks after discharge, i.e. complete resolution and residual lesions on CT.

### Statistical Analysis

We constructed the epidemic curve by date of illness onset, with interpretation of key dates relating to control measures. Data of cases with illness onset as of February 10 has been analyzed because we have found the obviously decrease of secondary infection identified after the formal announcement of the first-level public health emergency response of Xi’an. Case characteristics were described, including demographic characteristics, exposures, as well as clinical and imaging findings.

All statistical analyses were performed in the SPSS Statistics Software (version 22; IBM, New York, USA). Continuous variables were described using mean ± standard deviation and the categorical variables were described as the number and percentage of the total. The SPSS curve estimation module was constructed to estimate the total CT scores of lungs as a function of time. Proportions for categorical variables were compared using the Chi-square test or Fisher’s exact test. Mann-Whitney U test was used for comparing the continuous variables when the data unfit independent group *t* test. A p-value of <0.05 was considered statistically significant.

## Results

### Epidemiological and demographic characteristics

Of 104 confirmed cases, 99 patients were included and 5 people were excluded because they didn’t provide exposure histories (Fig. 1). The detailed characteristics of the study cohort were summarized in Table 1. Of the remaining 99 COVID-19 patients, 45 were first-generation, 36 were second-generation and 18 were third-generation infections. The third- and second-generation cases were significantly older than first-generation patients (mean±SD age, 43.13±14.96 years, 52.53±14.99 years vs 54.56±16.75 years; *p*<0.001). Besides, the second-and third-generation cases were more likely to have comorbidity, particular hypertension, than first-generation cases (11.1% of first-generation, 52.8% of second-generation vs. 55.6% of third-generation; *p*<0.001). As shown in Table 2, we examined data on exposures of secondary cases, 19 (35.2%) cases and 12 (22.2%) cases had a history of exposure to family or party including at least one first-generation patient. In Fig. 2, the development of the epidemic follows an exponential growth in cases, and two overlapping waves of transmission could be observed on the epidemic curve. The latter part of the curve indicated a decrease in the number of confirmed patients of secondary infections in Xi’an.

**Figure 1.**
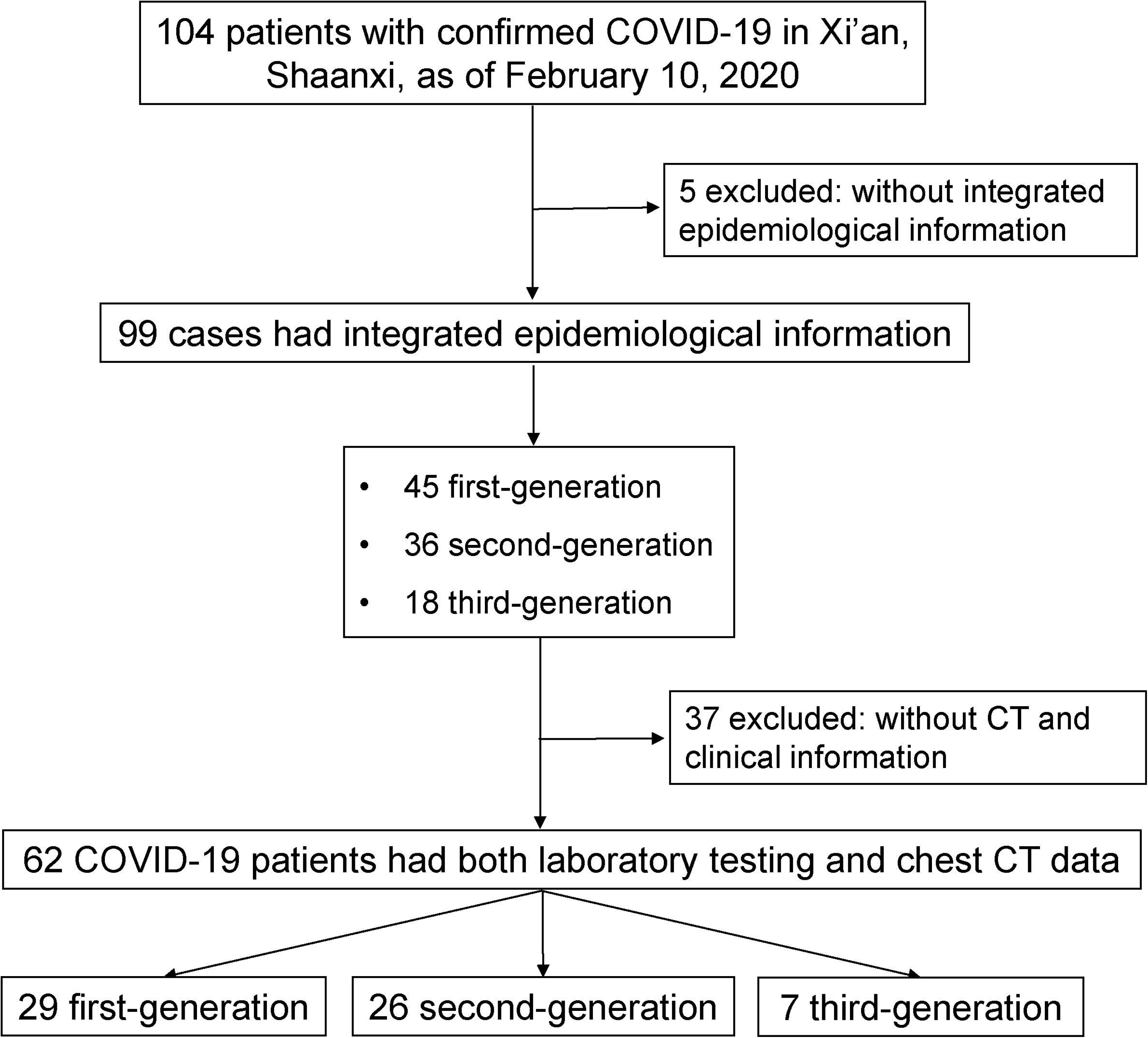
Flowchart of this study.

**Table 1.** Demographic characteristics of first-, second- and third-generation patients in Xi’an.

| Characteristic | Patients with COVID-19 |  |  | P value |
| --- | --- | --- | --- | --- |
|  | First-generation<br>(n=45) | Second-generation<br>(n=36) | Third-generation<br>(n=18) |  |
| Age group |  |  |  |  |
| 15-34 | 12(26.7%) | 6(16.7%) | 3(16.7%) | 0.286 |
| 35-44 | 15(33.3%) | 3(8.3%) | 1(5.6%) | **0.003** |
| ≥45 | 18(40.0%) | 27(75.0%) | 14(77.8%) | **0.001** |
| Male sex | 26(57.8%) | 16(44.4%) | 11(61.1%) | 0.380 |
| Comorbidity | 5(11.1%) | 19(52.8%) | 10(55.6%) | **<0.001** |
| Cardiovascular disease | 1(2.2%) | 4(11.1%) | 1(5.6%) | 0.242 |
| Hypertension | 4(8.9%) | 8(22.2%) | 5(27.8%) | 0.111 |
| Diabetes | 1(2.2%) | 1(2.8%) | 2(11.1%) | 0.240 |

Note: Unless otherwise indicated, data are reported as the number of patients, with percentages in parentheses.
The counting data were presented as count (percentage of the total).

**Figure 2.**
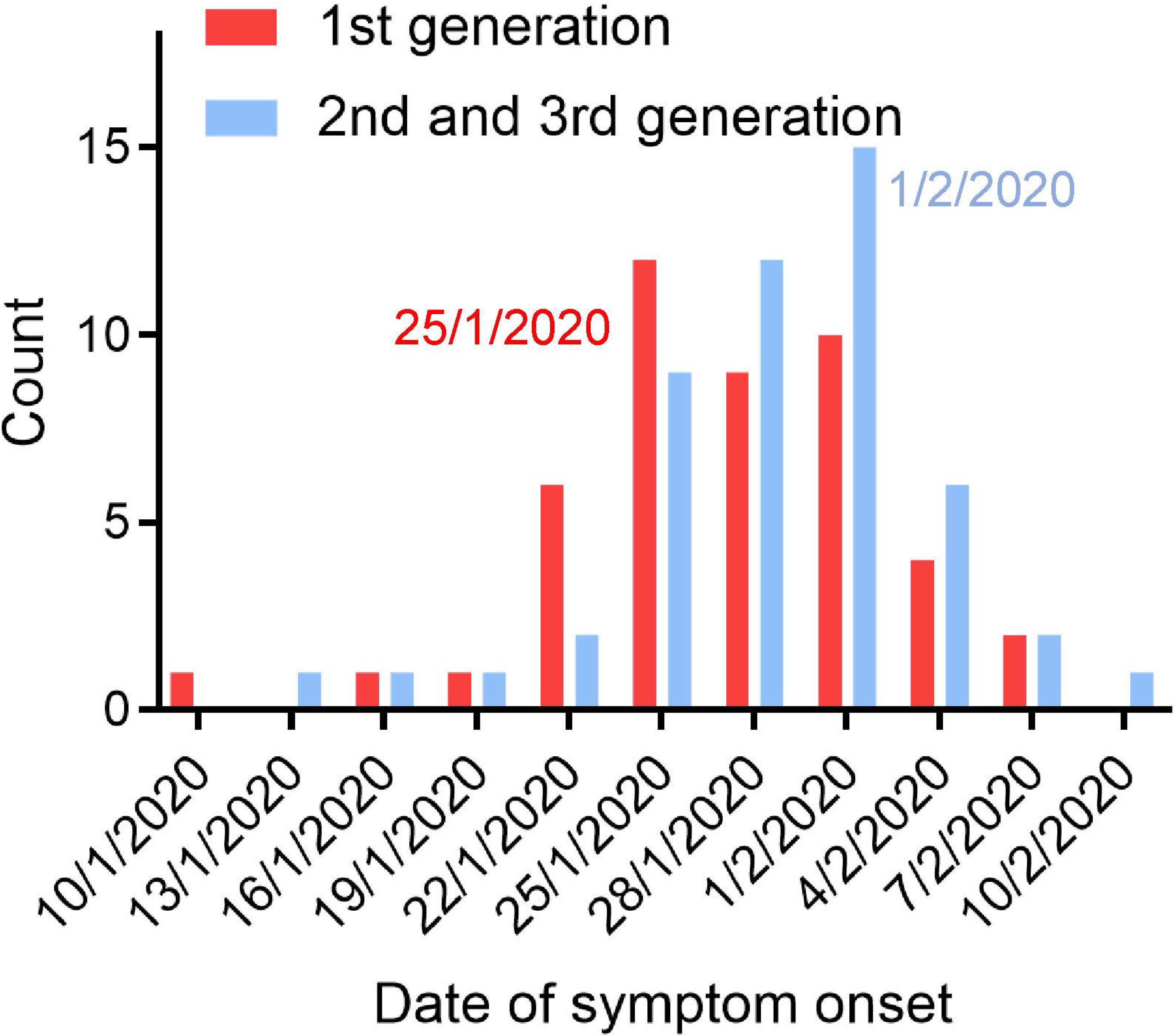
Epidemic curve showing the confirmed numbers of cases by generation (n=99) against time, as of February 10, 2020.

**Table 2.** Exposure history of secondary generation patients in Xi’an.

| Exposed at least once in | 2<sup>nd</sup> and 3<sup>rd</sup> generations (n=54) |
| --- | --- |
| Party | 12(22.2%) |
| Shopping mall | 9(16.7%) |
| Hospital | 3(5.6%) |
| Family exposure | 19(35.2%) |
| Unkown exposure | 11(20.4%) |

### Clinical and laboratory features at presentation

After exclusion of the cases without laboratory-testing data and chest CT, Table 3 shows the characteristics of 62 patients from three hospital in Xi’an including 29 first-generation, 26 second-generation and 7 third-generation cases. The detailed characteristics of patients in this study were summarized in Table 3. Of the 62 cases, the age of multiple generations shows significantly differences (42.14±14.96, 50.50±15.97 vs 57.86±16.36, *p*=0.030). Hypertension (6.9% of first-generation, 23.1% of second-generation vs. 28.6% of third-generation infections, *p*=0.138) was the most common comorbidity in all of multiple generation patients.

**Table 3.**
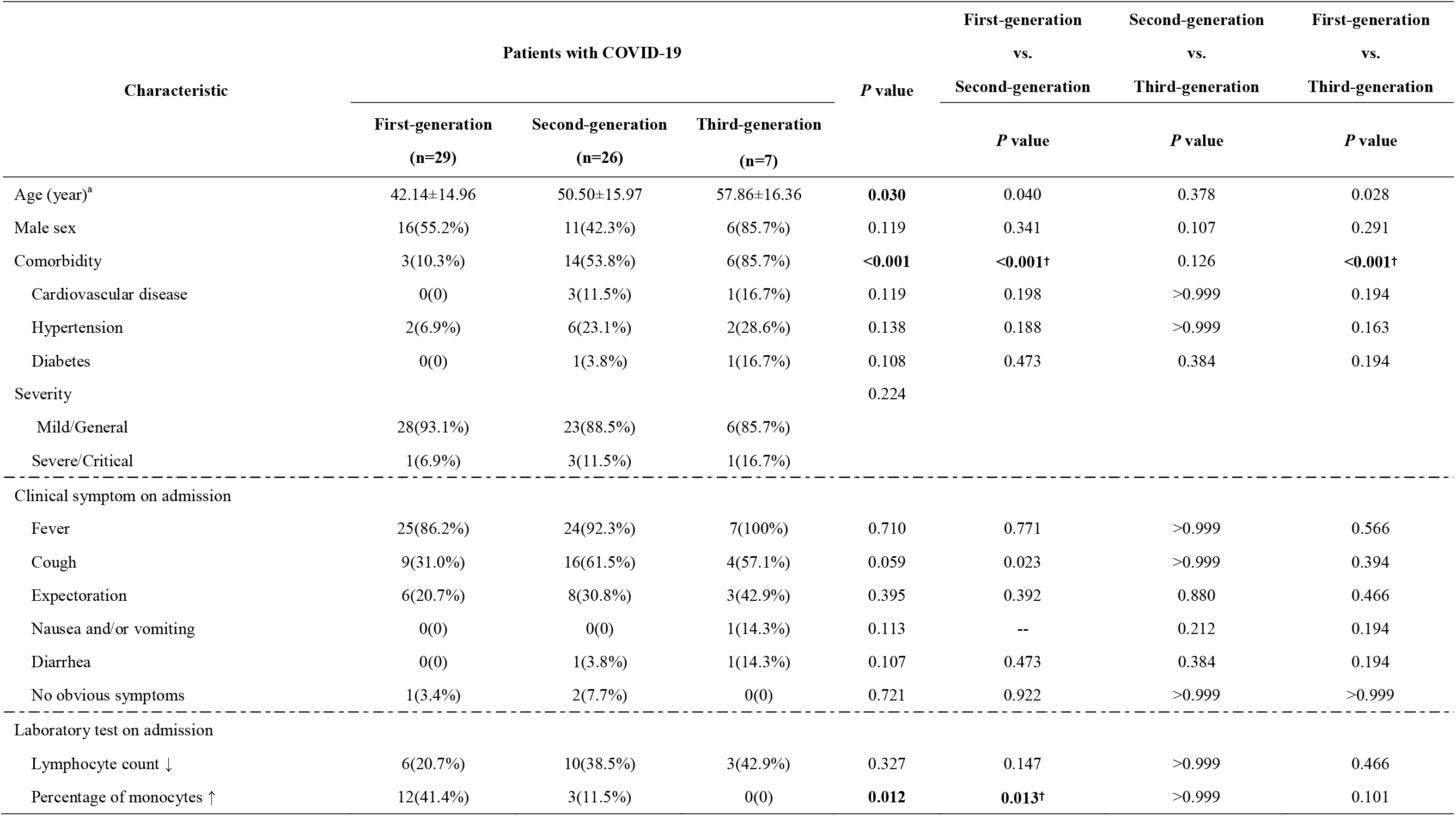
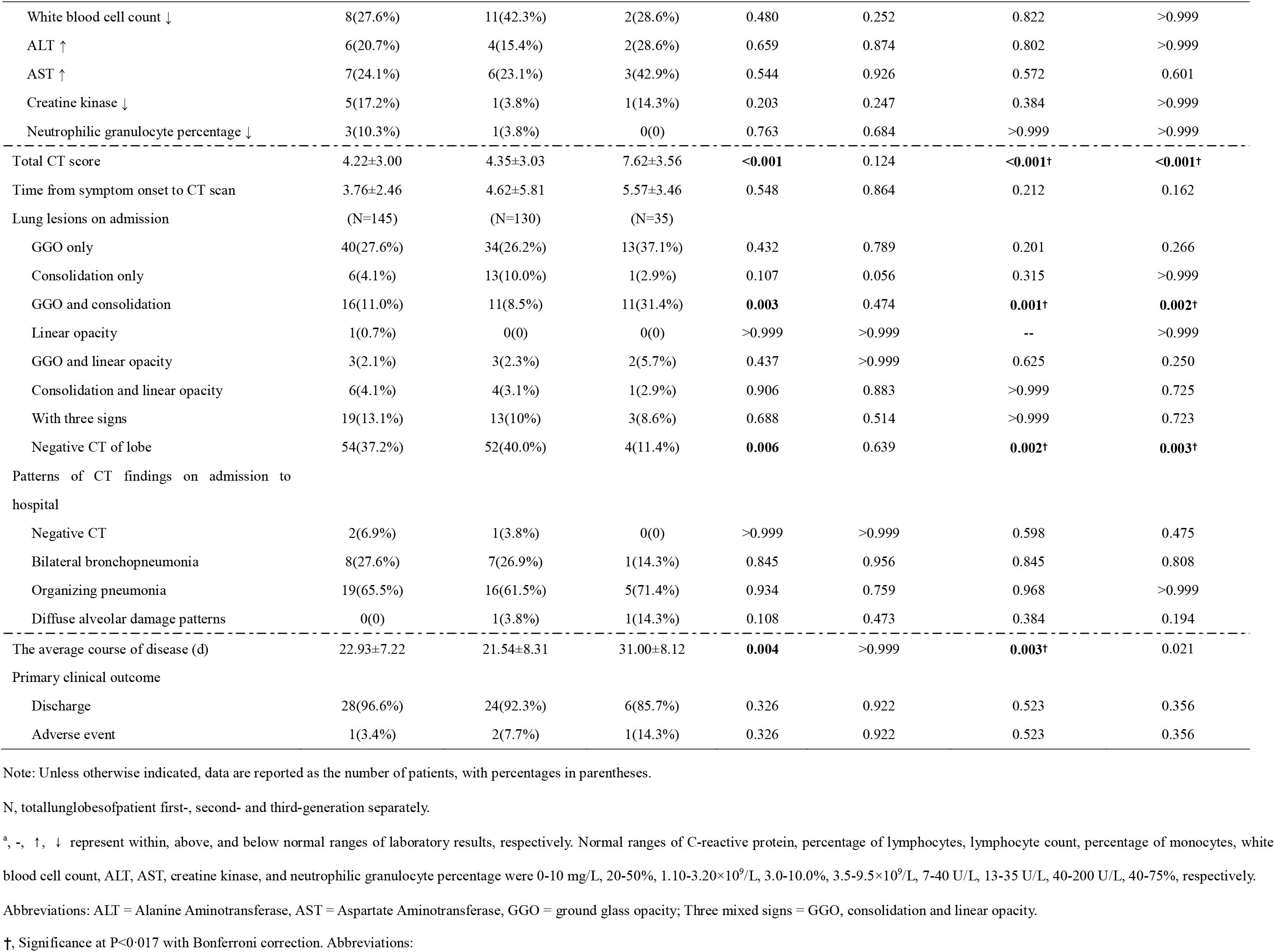
Characteristics of first-, second- and third-generation patients with COVID-19 infection.

The clinical characteristics and laboratory findings of the 62 cases by group are summarized in Table 3. Fever (86.2% of first-generation, 92.3% of second-generation vs. 100% of third-generation infections) was the most common symptom at onset. The initial laboratory test showed no significant difference in three groups except that percentage of monocytes increase was more significant in first-generation (*p*=0.012).

### CT characteristics at presentation

All 62 patients underwent initial CT average 3.9 days (range, 1-12 days) after onset of symptoms. Based on quadratic curve fitting between total CT score of pulmonary lesions and disease course in patients of three generations, the total CT score peaked at 10-15 days (10 days of first generation, 15 days of second-generation and 13 days of third generation infections) after the onset of initial symptoms and then gradually decreased (Fig. 3). As shown in Fig. 3 and Table 3, there was significantly more serious pulmonary infection in third-generation infections with higher total CT score than the first-and second-generation patients (*p*<0.001). The period between the onset of initial symptoms and the time of first CT scan showed no significant differences between three groups (*p*>0.05). Furthermore, the average courses of the disease in third-generation infections has obviously extension to 31.00±8.12 days (*p*=0.004). Moreover, there were no significant differences of the pulmonary sequelae in three generation patients (Table 3).

**Figure 3.**
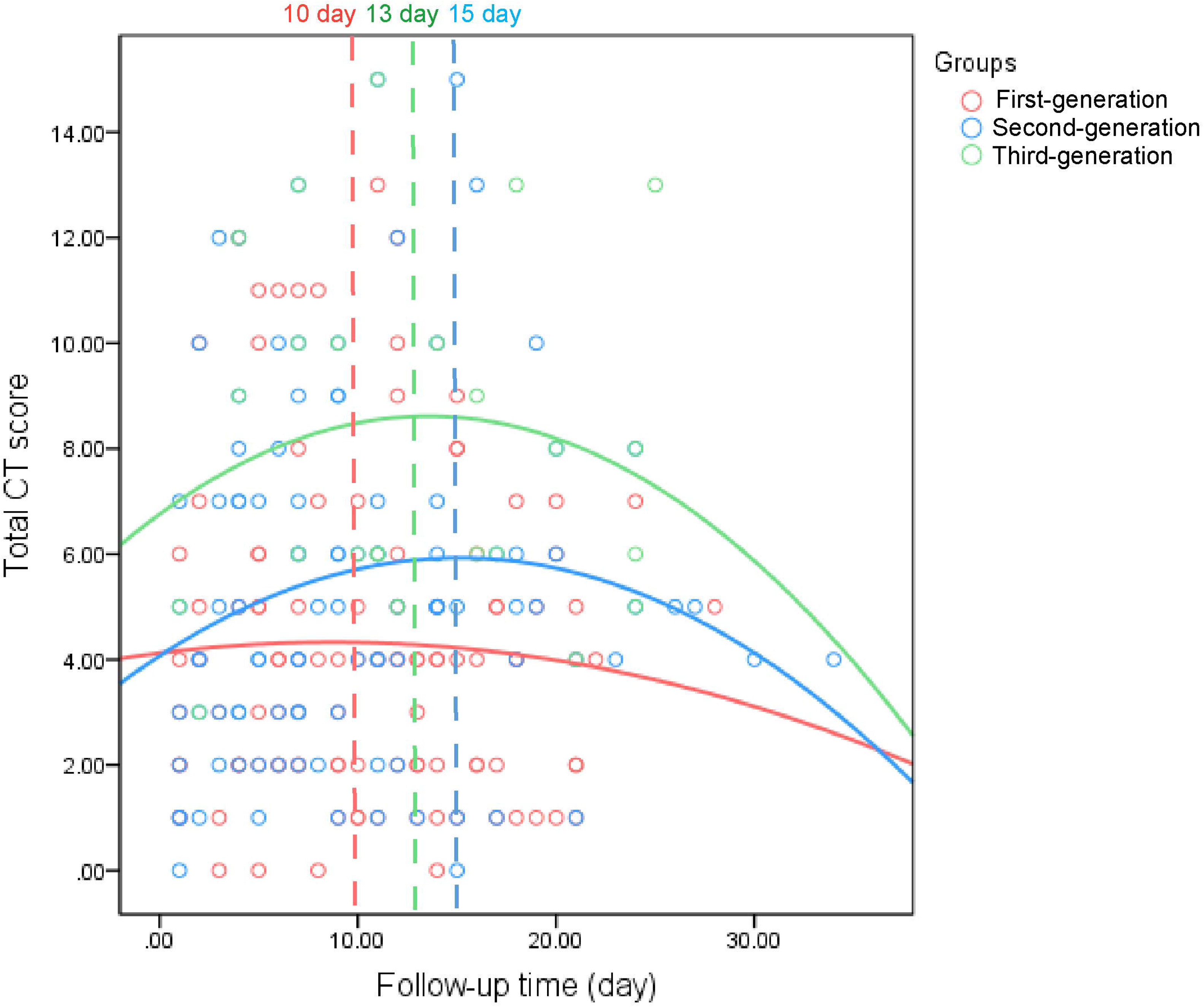
Dynamic changes of total CT score in pulmonary lesions from time onset of initial symptoms in first- and second-generation. (Curve fitting equation: first-generation, y=-0.0027*x^2^+0.05*x+4.12, R^2^=0.002; second-generation, y=-0.0082*x^2^+0.25*x +4.07, R^2^=0.029; third-generation, y=-0.01*x2+0.27*x+6.76, R2=0.024; x=time from the initial symptoms appear, y=total CT score of the pulmonary lesions).

GGO only (27.6% of first-generation, 28.5% of second-generation vs. 37.1% of third-generation infections, p=0.432) were the most common CT findings in the initial period in all of the three generation cases with C0VID-2019 infection. However, in third-generation cases, GGO and consolidation (11.0% of first-generation, 8.5% of second-generation vs. 31.4% of third-generation infections, *p*=0.003) was more familiar, with less negative CT (37.2% of first-generation, 40.0% of second-generation vs. 11.4% of third-generation infections, *p*=0.006). As shown in Fig. 4, a categorization of radiological patterns was performed by detailing the extent of lung injury in COVID-19. In all of three generations, organizing pneumonia (65.5% of first-generation, 61.5% of second-generation vs. 71.4% of third-generation infections, *p*=0.934) were the most frequent CT patterns in COVID-19. Remarkably, diffuse alveolar damage patterns only disappeared in the first-generation infections in our cohort (3.8% of second-generation vs. 14.3% of third-generation infections, *p*=0.108).

**Figure 4.**
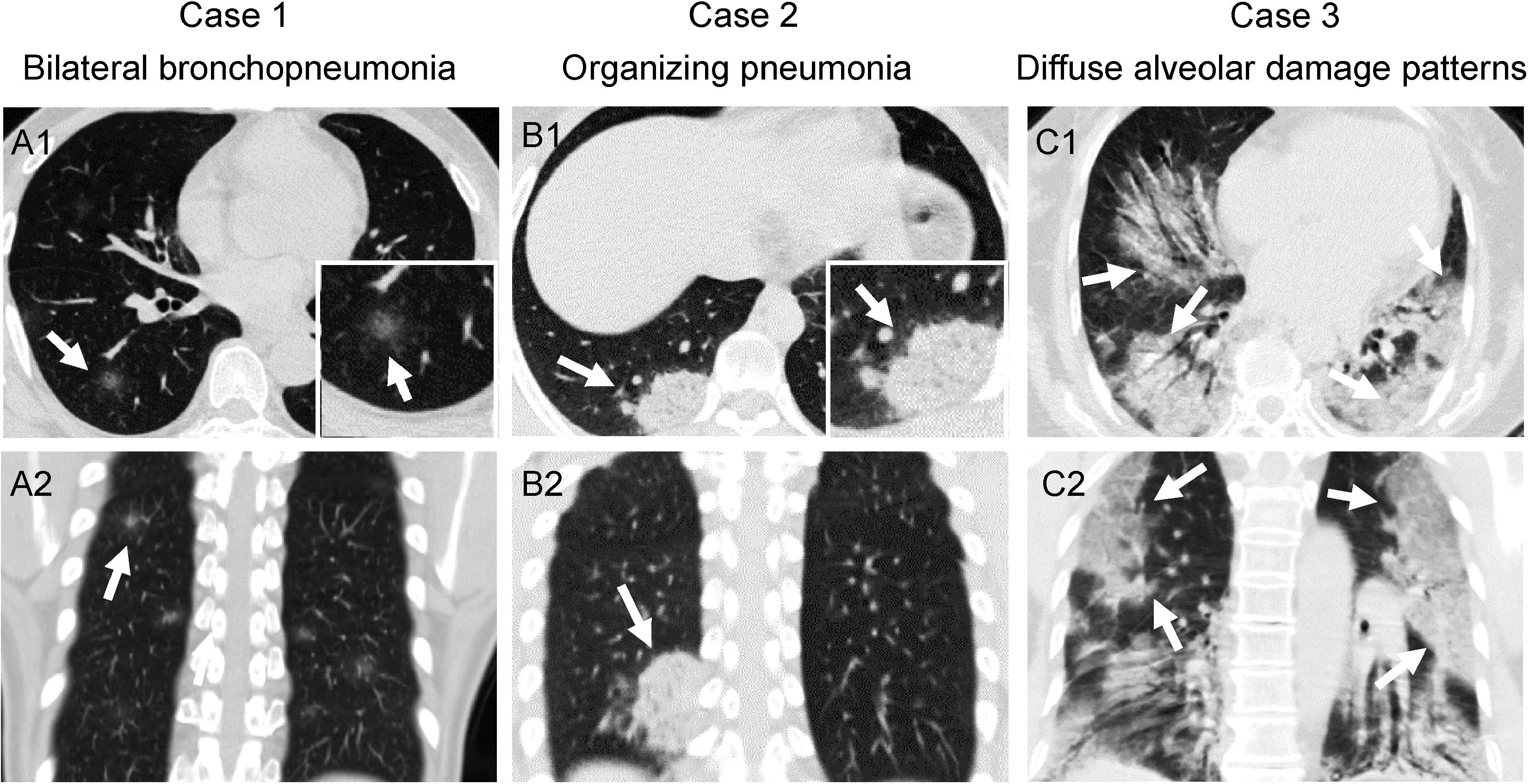
Typical chest CT examples of three patterns (arrows) of COVID-19 penumonia. Case 1.55-year-old female of the first-generation infection. A1, A2 initial CT scans obtained on day 4 after the onset of symptom showed bilateral bronchopneumonia which was defined as mild infection; Case 2. 22-year-old female of the first-generation infection. B1, B2 initial CT scans obtained on day 3 after the onset of symptom showed organizing pneumonia; Case 3. 66-year-old female of the second-generation infection. C1, C2 initial CT scans obtained on day 3 after the onset of symptom showed diffuse alveolar damage pattern which was defined as critical infection.

## Discussion

Detailed investigation of transmission chains of imported COVID-19 pneumonia is crucial for understanding which control measures are necessary and which of them need to be reinforced. The age effect is obvious in secondary generation patients. The COVID-19 virus infects people of all ages, while evidence to date suggests that older people with underlying medical conditions are at higher risk of getting infections (3). The infection of COVID-19 coincides with the peak of the Chinese Spring Festival travel rush so that it became a serious public health concern. Beginning in December 2019, Chinese health authorities have been closely monitoring the cluster of COVID-19 pneumonia cases in Wuhan. On January 23, 2020 at 10:00, travel restrictions was executed in Wuhan. From the analysis of data, we find that secondary infections of first-generation increased fast in Xi’an before 25 January, 2020, but quickly decreased during about ten days due to the interruption of transmission by travel restrictions (17). Similarly, Shaanxi province has activated first-level public health emergency response since 25 January, 2020 (18). A series of measures which aim to restrict social contact such as home quarantine, traffic control and suspension of public places was announced and worked. Therefore, second- and third-generation infections increased to the peak in early February and followed with reduction during ten days. These results demonstrated that subsequent measures in Wuhan and Xi’an have reduce transmissibility effectively, such as the traffic blockage and population quarantine.

Transmission at household and parties were found to be two most important cause until controls were put in place, due to the courtesy of visiting relatives before the Spring Festival. Hospital infection had a minor role in this outbreak attributing to the efficient assessment and management to health workers exposure risk in hospital. Currently, WHO offered guidance on quarantine to member states, which involves the restriction of movement or separation of healthy individuals who may have been exposed to the virus from the rest of the population, for infection prevention and control measures (19). We also suggest keeping the following for individual, particular the vulnerable elderly population, acts to cooperate with the prevention and control of the virus spread: minimize outgoing, avoiding dining together, wash hands as often as possible and wear masks.

In our study, we focused on whether there are differences in clinical characteristics and chest CT signs among multiple generation patients with COVID-19 infection. Patients with COVID-19 infection may present primarily with fever and cough in all three generation cases, which is consistent with previous research (20). There was no difference in clinical symptoms among patients of different generations. At the time of admission, some of the three generations of patients had leukopenia and hypolymphocytes. The incidence of increased percentage of monocytes in first-generation cases is higher than in second- and third-generation patients. Previous investigations suggested that lymphopenia might be a key factor related to the severity and lethality of COVID-19 infection, while the monocytes was unusually noted for this disease (21). Monocyte is considered to be the precursors of macrophages. It has the functions of phagocytosis and elimination of pathogens, and participates in immune reactions. After phagocytosis of antigens, it carries the antigenic determinants to lymphocytes to induce specific immune responses of lymphocytes (22). Thus, we postulated that there are differences in body inflammation and immune status between the three generations, which may be related to the patient’s individual heterogeneity.

Chest CT is a key component of the diagnostic for COVID-19 infection not only for early screening of suspected cases but also for severe complications and prognosis prediction (23-25). Then we provide an initial assessment of the CT findings and time course of severity of lung changes among three generations patients. In all of the three generations cases with COVID-2019 infection, the most common pulmonary CT findings were GGO only. This was consisted with prior reports (15). GGO and consolidation only was more prevalent in the third-generation patients in the initial CT imaging, which demonstrated aggravated infections on admission to hospital. These findings represent that pneumonia of third-generation cases were more severe and more complicated than the second-generation and first-generation cases. The dynamic changing curve of total CT score in pulmonary lesions showed that severity and number of lesions increased in the first 10-15 days in all of the three generations patients, followed by a gradual decrease, which was accord with the progress of this disease (26). Nevertheless, the third-generation patients had obviously higher total CT score at the same timestamp. The reason may be related to more comorbidities in second- and third-generation cases. Meanwhile, third-generation cases had extended course of the disease than other two generation cases. This indicated that comorbidity and severity of pneumonia in CT findings might explain the longer course of the disease in third-generation cases.

Viruses in the same family have the same pathological mechanism and will also lead to similar imaging findings (27). Furthermore, recent investigations characterized predominant radiological manifestations of COVID-19 pneumonia, including GGO, consolidative pulmonary opacities, peripheral and diffuse distribution, bilateral involvement, which overlapped with various virus pneumonia (28). Lee put forward the pattern approach of classifying the viral pneumonia into three patterns: bronchopneumonia, organizing pneumonia, and diffuse alveolar damage patterns, which had been recognized and characterized thoroughly in previous histopathological and radiological studies (16, 29). This classification may help stratify viral pneumonic patients in terms of their prognosis. Prior studies retrospectively assessed the relationship of these CT findings with clinical outcome and they concluded that patients presenting with diffuse alveolar damage pattern were prone to show more severe clinical course and adverse outcome, while patients with bronchopneumonia and organizing pneumonia pattern tended to present the mild illness and longer survival (30, 31). In our study, we found that organizing pneumonia was the major pattern whether in the first-, second- or third-generation cases. However, there was no statistical difference in the type of pneumonia between the three groups. This is consistent with the clinical diagnostic classification of our cases. The evidence above seems to explain why there were no significant differences in outcomes, adverse events, and post-discharge pulmonary sequelea at follow-up between the three generations, although the severity of lung lesions is different. Considering that there are no obvious differences of clinical characteristics in multiple generations, we speculate that there is no obvious virus mutation spread among three generations in our cases.

The several limitations include in this study. First, the samples we investigated is small, particularly for patients defined as sever/critical. As more data become available in other locations, it will be possible to promote this investigation. Second, we compared epidemiological, demographic, clinical and radiological characteristics in the multiple generation infections in Xi’an, which does not account for potential geographic discrepancy.

In conclusion, the government interventions against COVID-19 have been successfully used to stop the epidemic in Xi’an. Combining clinical features and chest CT patterns, our study demonstrated similar changes in three generations of cases. Older people with underlying medical conditions are at higher risk of getting secondary infections and longer duration. Measures for public and individuals to reduce or prevent transmission should be reinforced in populations at risk in the next stage.

## Main Points

1. Measures to reduce or prevent transmission should be reinforce in older people with comorbidity who are at higher risk of getting COVID-19 infections in second- and third-generation.
2. There are no obvious differences of clinical characteristics in multiple generations of COVID-19 infections.
3. Organizing pneumonia is a pattern of CT findings in all of three generations with COVID-19 infection.

## Data Availability

Some or all data, models, or code generated or used during the study are available from the corresponding author by request.

## Acknowledgments

This study was supported by “Xi’an Special Science and Technology Projects on Prevention and Treatment of Novel Coronavirus Pneumonia Emergency” (Grant Number: 20200005YX005).

